# Leptin augmented model to include the role of obesity in insulin-glucose regulatory system for T2DM subjects

**DOI:** 10.1101/2024.06.18.24309097

**Authors:** Manoja Rajalakshmi Aravindakshan, Devleena Ghosh, Chittaranjan Mandal, Jit Sarkar, Sujay K Maity, Partha Chakrabarti

## Abstract

Leptin is a fat cell-derived hormone involved in satiety and body weight regulation. It also plays a critical regulatory role in the insulin-glucose regulatory system by modulating glucose metabolism and energy homeostasis. However, existing insulin-glucose models often fail to consider the impact of body weight indicators mainly body mass index (BMI) and plasma leptin. To address this limitation, we propose augmenting the ordinary differential equations (ODE) of the Oral Minimal Model (OMM) with an additional equation, incorporating leptin as well as supplementary terms and parameters. By estimating the model parameters, the model behaviour is aligned with the observed data of glucose, insulin and leptin for individuals with type 2 diabetes mellitus (T2DM). Based on model behaviour, revised indices formulated from Oral Glucose Tolerance Test (OGTT) data by including BMI and fasting leptin values are found to have a better correlation with existing indices. Additionally, parameter sensitivity analysis is performed to investigate the influence of the model parameters on the observed variables. Validation of the augmented model with clinical data (without leptin) demonstrates a superior fit to glucose and insulin data compared to the base model. This model emphasizes the intricate associations between leptin, glucose, and insulin concentrations with a potential for developing targeted interventions and therapies for T2DM. Notably, this manuscript introduces the first ODE-based model that incorporates leptin and BMI in the insulin-glucose pathway.

## 1 Introduction

Insulin glucose (IG) regulatory system is a biochemical system that helps to maintain a steady glucose level in the blood. The main source of glucose is from meal ingestion (exogenous input) and excess glucose is converted to glycogen by the liver and stored there. When the level of glucose falls in the blood (due to exercise or a long gap after the last meal), pancreatic *α*-cells secrete glucagon to release glucose (glycogenolysis) from the liver through the breakdown of glycogen until the glucose level rises to normal. When the level of glucose rises in the blood (after a meal), pancreatic *β*-cells secrete insulin to trigger the uptake of glucose by the peripheral tissue cells in the body until the glucose level falls back to normal. Type-2 diabetes mellitus (T2DM) occurs when the body becomes resistant to the effects of insulin and the pancreas fails to produce enough insulin to compensate for this resistance [1]. Excess body fat (obesity), particularly abdominal or visceral fat, is associated with insulin resistance (IR), a condition in which the cells of the body become less responsive to insulin. This reduced sensitivity to insulin leads to elevated blood glucose levels resulting in the characteristic symptoms and complications of T2DM [1]. Obesity-induced IR is followed by *β*-cell dysfunction leading to T2DM, however, the contribution of obesity and *β*-cell dysfunction in T2DM development vary significantly among the non-obese and obese population [2, 3].

Obesity is a major risk factor for the development of T2DM[4]. Leptin, a hormone secreted by fat cells, plays a crucial role in regulating body weight and energy balance. Leptin acts on the hypothalamus to suppress appetite and increase energy expenditure [5]. Additionally, leptin has various secondary functions in peripheral tissues, wherein it influences insulin level and glucose metabolism in the body. In the relationship analysis between leptin and body mass, serum leptin concentration is found to have a direct positive association with body mass index (BMI) [6]

There are several theoretical works [7, 8] that emphasize the role of leptin in IR and its importance in T2DM. Mathematical models including leptin in diet and type 1 diabetes [9–11] exist with numerical simulation results, however, we aim for a model for T2DM that can capture leptin behavior using OGTT data. Our objective is to not only conceptualize such a model but to substantiate its real-world relevance through rigorous validation using clinical data. This is achieved by augmenting the Oral Minimal Model (OMM) proposed by Dallaman et al. [12, 13] to include the role of leptin and BMI. Our model aims to incorporate the regulatory influence of leptin on insulin-glucose dynamics, thereby improving the existing model fitting of glucose and insulin data and accurately capturing the leptin data. Consistent with the OMM approach, the current model uses clinical data derived from the Oral Glucose Tolerance Test (OGTT) for parameter estimation and analysis.

OGTT is a frequently used diagnostic test in which the subject’s fasting blood glucose levels are measured, followed by the ingestion of a specific amount of glucose dissolved in water, and subsequently, blood is collected at regular intervals to measure glucose and insulin levels at varying intervals [14]. It is a highly sensitive test used for screening and diagnosis of T2DM. The dataset used for modeling in the current work consists of data of individuals who have undergone a 2-hour OGTT. In addition to insulin and glucose, leptin concentrations are also taken at various time points for this study. OGTT indices (derived from OGTT data) are quantitative measures that provide information about the subject’s insulin sensitivity, *β*-cell function, and overall glucose regulation in response to a glucose load. The current work attempts to suggest modifications and enhancements to selected OGTT indices.

One of the crucial aspects of modeling is to understand how parameter values change with model output. The parameter variations affect the output of a model when dealing with critical data specific to biochemical pathways. The interpretation of parameter values in these pathways is important to understand the relationship to their physiological properties. Sensitivity analysis on the augmented model is also conducted to explore the behaviour of model parameters, providing inference on the model’s outcomes. Additionally, validation of the model is performed using a different dataset, as detailed in the subsequent sections. The present study introduces a novel approach to modelling the interaction between leptin and the insulin-glucose pathway, which has not been previously explored in the existing literature. The following sections describe the background, augmented model and parameter estimation followed by results and discussion.

## 2 Background

ODE modelling started with the *minimal model* for modelling the intravenous glucose tolerance test (IVGTT). Bergman’s minimal model is a well-accepted coupled model from the original model [15]. The simplest form of OMM [12] as ordinary differential equations (ODE) are represented in Eqn. 1:

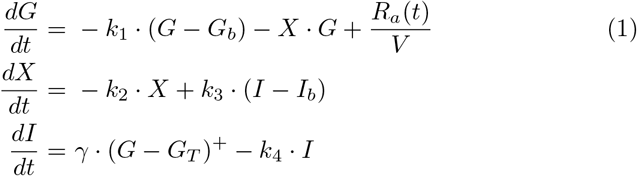

where *G, I* denotes blood glucose concentration and blood insulin concentration (suffix *b* denotes basal values) at time *t*, respectively. Here, *X* is an auxiliary variable to model the time delay in insulin-dependent glucose uptake activity. The initial values of G and I are taken at the time when the glucose is orally ingested. The initial value of *X* is taken as 0 and (*G*−*G*_*T*_)^+^ = (*G*−*G*_*T*_), if *G > G*_*T*_ and 0 otherwise. *R*_*a*_ is the rate of absorption of ingested glucose; *V* is the volume of distribution; *k*_1_ is the fractional glucose effectiveness i.e., the ability to promote glucose disposal and inhibit glucose production, *k*_2_ is the insulin-dependent increase in glucose uptake ability in tissue per unit of insulin concentration above *I*_*b*_, *k*_3_ is a scale factor for the amplitude of insulin action and *k*_4_ is the first-order decay rate for insulin in plasma. There are several complex IG models that have evolved over time ([16, 17]). The OMM is based on the Bergman minimal model [15] in which the glucose analysis is done using IVGTT. The variable *X* in the minimal model could be more accurately understood as representing the incremental insulin response within the interstitial space in response to a bolus stimulus, rather than being directly proportional to the interstitial insulin level, as it was conventionally assumed [18]. A quantitative physiological model for insulin glucose regulatory system will be more suitable for epidemiological studies and this is achieved in the OMM developed by Dallaman et al. [12] based on OGTT. The model presented in this paper is based on the well-cited work of Dallaman et.al.[12, 19] on the OMM.

The rate of appearance of oral glucose in plasma is coupled with minimal model of glucose kinetics through the parameter *R*_a_. The Model-2 in the work of Man et al.[13], hereafter referred to as R_*a*_ model, is used in the current work with a modification, which is in accordance with the non-linear gastric emptying of glucose (liquid phase). The solid phase glucose compartment from the stomach is removed as it deals with a grinding rate parameter which is not relevant to our study as glucose solution is orally ingested. The R_a_ model equations are given as follows (Eqn. 2):

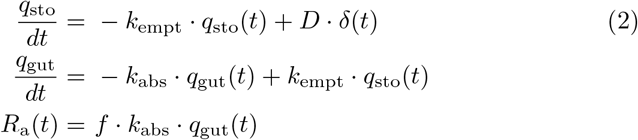

where *q*_sto_ is the amount of glucose in the stomach, *δ*(*t*) is the impulse function, *D* is the amount of ingested glucose, *q*_gut_ is the glucose mass in the intestine, *k*_empt_ is the rate of gastric emptying and is a function of *q*_*sto*_ [13], *k*_abs_ is the rate constant of intestinal absorption, and *f* is the fraction of the intestinal absorption which actually appears in plasma. This model with ODEs of OMM (Eqn. 1) is considered as *base* model from hereon.

By efficiently integrating BMI and the adipokine leptin into the existing OMM, their associations with insulin and glucose concentrations can be effectively modelled. Leptin is associated with insulin resistance and is positively correlated with BMI [20]. Leptin decreases insulin secretion to a certain extent [21] and also exhibits a negative correlation with fasting plasma glucose and postprandial glucose after two hours. Leptin also decreases hepatic glucose production, increases insulin sensitivity, and decreases glucagon levels. This inhibitory action of leptin is modelled using enzyme kinetics in this work. Insulin, in turn, also plays a role in stimulating leptin production and secretion in the adipose tissue [7]. Leptin deficiency results in impaired glucose metabolism and prolonged insulin resistance [8].

The commonly used indices to access beta cell function and insulin resistance are briefly explained here. HOMA-B (Homeostatic Model Assessment of Beta-Cell Function) is a mathematical model used to estimate *β*-cell function based on fasting glucose and fasting insulin levels, particularly in the context of insulin resistance and T2DM. The formula for calculating HOMA-B is 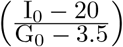, where I_0_ is fasting insulin (in *µU/mL*) and G_0_ is fasting glucose (in *mmol/L*) values. HOMA2 is an improvement over the original HOMA model, as it considers glucose dynamics across populations and different physiological states. Several software tools that accurately calculate HOMA2 values use advanced algorithms as in the HOMA calculator [22] provided by the diabetes trials unit at the University of Oxford. HOMA-IR (Homeostatic Model Assessment of Insulin Resistance) is a widely used method for estimating insulin resistance, which is a key component of various metabolic disorders, including T2DM. The formula for calculating HOMA-IR is 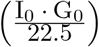, where I_0_ is in *µU/mL* and G_0_ is in *mmol/L*.

The insulin sensitivity index (ISI) is derived from the concept that insulin sensitivity can be assessed by evaluating the ability of insulin to regulate glucose levels. The formula for calculating ISI is 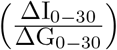, where Δ is difference in fasting and 30 min values of insulin(I) and glucose(G). Higher ISI values indicate greater insulin sensitivity, meaning that the cells in the body are more responsive to insulin and effectively utilize glucose. The oral disposition index, DI_o_ [23] calculated as 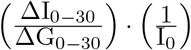, where I_0_ is in *pmol/L* and G_0_ is in *mmol/L*, is commonly used as a measure of *β*-cell function.

The idea of parameter estimation is to find unknown parameters in a computational model which may describe the given biophysical pathway or phenomenon. These unknown parameters are estimated using experimental data which are collected from well-defined and standard conditions by minimizing the error between the model simulations results and experimentally known data [24]. This way the behaviour of the model is captured effectively. The challenge is that no single optimal estimation technique exists for all models. Many different estimation methods have been developed so far to determine the best strategy for a given problem. The commonly used parameter estimation methods include maximum likelihood estimation [25] and Nelder Mead optimization [26]. Nelder Mead method is a numerical method in non-linear optimisation problems to find the minima of objective functions. Least square estimation is used in regression models and maximum likelihood estimation is used in statistical models [27]. In addition evolutionary methods[28] namely genetic algorithms and particle swarm optimization are other efficient parameter estimation methods used in non-linear dynamic models.

Our aim is to develop an underlying well-established model that includes the role of BMI and leptin by estimating the set of parameters that are significant.

## 3 Augmented model

Leptin level plays a major role in glucose homeostasis and is positively correlated with adiposity. In the presence of high blood glucose levels, leptin enhances tissue glucose uptake, independent of insulin, and reduces hepatic glucose production [7]. Leptin has glucose-lowering effects via an insulin-independent mechanism which normalizes hepatic glucose production and increases glucose uptake in peripheral tissues including adipose tissue, muscle and heart [29]. Insulin promotes adipogenesis (formation of fat cells from pre-adipocytes) and increases body fat mass [30]. It also stimulates the production and release of leptin, which acts centrally to decrease food intake and boost energy expenditure. Leptin, in turn, inhibits insulin secretion through both central and direct effects on *β*-cells. As plasma leptin level is directly linked to body fat mass, higher adiposity raises plasma leptin, thereby reducing insulin production and promoting further increasing fat mass [31] (Fig. 5). In this way, both leptin and insulin regulate each other, sharing the control of food intake and metabolism [7].

**Fig. 1:**
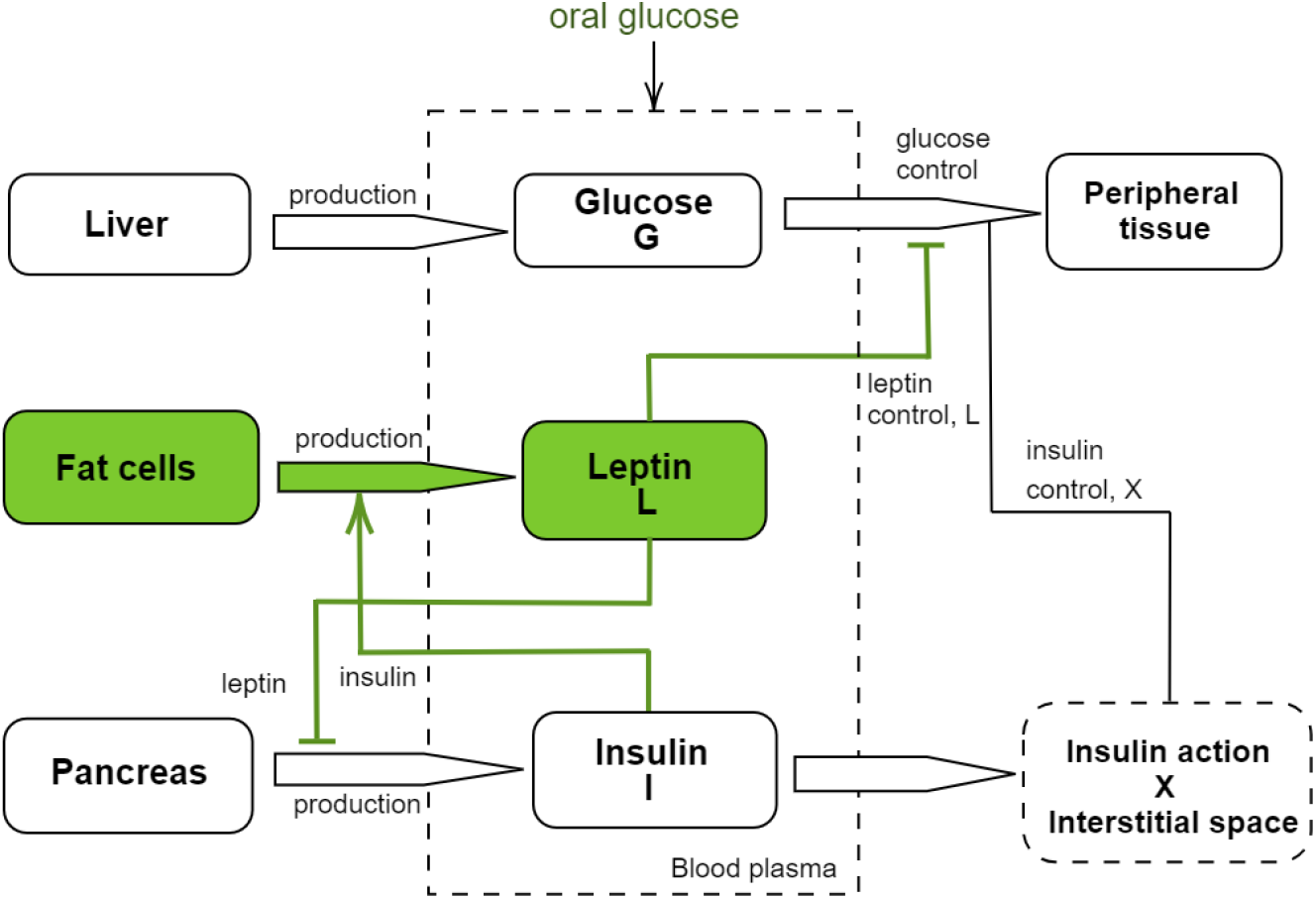
*Leptin* model: The augmentations to the model are visually highlighted using the colour green. Leptin has glucose-lowering effects independent of insulin. Insulin increases the production of leptin by adipose tissue (fat cells). Leptin feeds back to reduce insulin secretion.

**Fig. 2:**
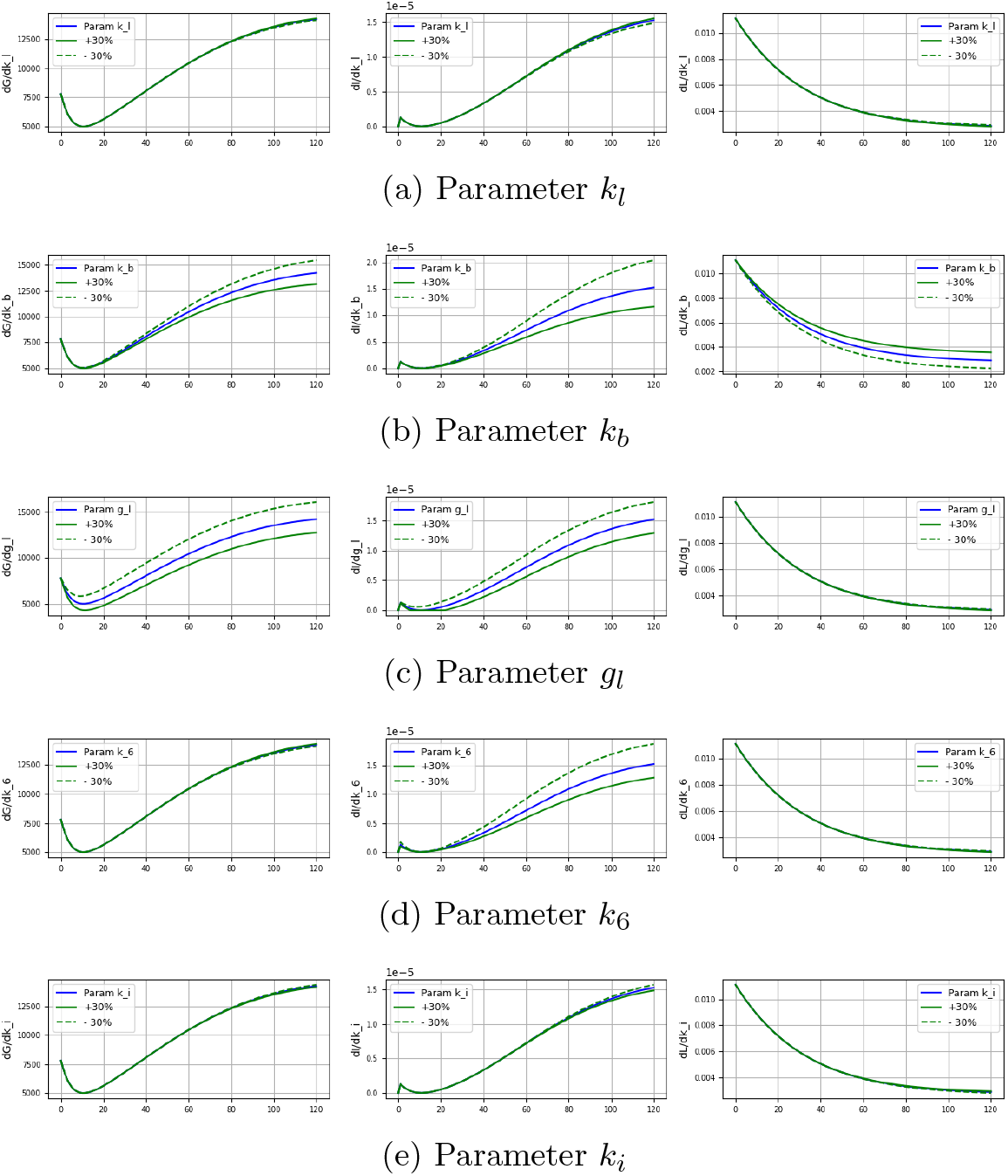
Sensitivity analysis of parameters with ± 30% variation wrt glucose, insulin and leptin

BMI which is a measure of body fat, is based on the weight and height of an adult. Adipose tissue produces leptin in quantities proportional to its mass and thereby regulates body weight. Moreover since leptin level increases with BMI [32] we have modelled the regulatory role of leptin and BMI in the IG regulatory system by augmenting the oral minimal model.

The augmentations carried out in the OMM are as follows:

- leptin-induced glucose absorption in peripheral tissues independent of insulin[5, 33],
- leptin-induced inhibition of insulin secretion [7],
- leptin production from adipose tissue modelled through BMI. This production can be dependent and independent of insulin [6, 7].

For accommodating leptin concentration, one additional differential equation (DE, Eqn. 7) is added and additional terms (*T*_1_ − *T*_6_) in the DEs for glucose and insulin concentrations (Eqns. 3, 5) are added. Also, an expression (Eqn. 6) for capturing the effect of BMI is introduced. The augmented model equations are given as follows:

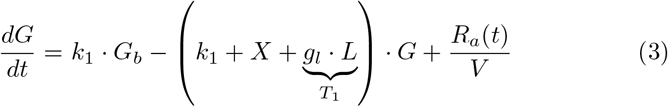

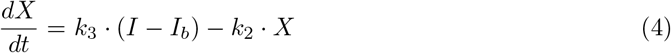

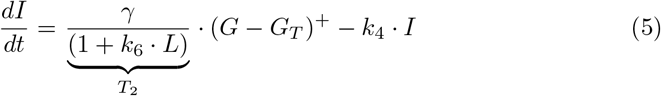

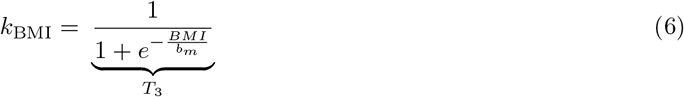

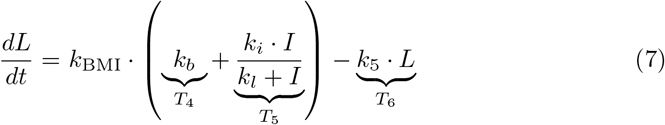

where *L* is the leptin concentration in the blood, *BMI* is the body mass index of the subject, *g*_*l*_ denotes the ability of leptin to lower blood glucose levels, *k*_6_ denotes the inhibitory action of leptin on insulin, *b*_*m*_ is a scaling factor that regulates the effect of BMI, *k*_*b*_ is the BMI contribution factor to leptin production, *k*_*i*_ denotes leptin production based on insulin, *k*_*l*_ denotes the ability of insulin to stimulate leptin production and *k*_5_ is the leptin degradation rate.

The ability of leptin to independently lower blood glucose levels is modelled as *T*_1_ in Eqn. 3. The effect is coupled with glucose as there exists a co-regulation between leptin and glucose via insulin. As leptin inhibits insulin production [21], this behaviour is modelled assuming reversible competitive inhibition [34] as *T*_2_ in Eqn. 5. The role of BMI in leptin production is modelled empirically as a logistic function (Eqn. 6) to incorporate the behaviour of leptin for the BMI range of [18, 40]; the parameters are determined through regression using dataset-1. An additional equation for leptin concentration (Eqn. 7) is added by introducing the effect of BMI and insulin in leptin production and a decay term to model the degradation of leptin over time. The basal level of leptin is handled in the model using the term *T*_4_ in the Eqn. 7 for leptin *k*_BMI_ is a function of BMI that regulates the effect of BMI in leptin concentration with a scaling parameter *b*_*m*_. As leptin is mainly produced from body fat this effect is introduced by the term *k*_BMI_ · *k*_*b*_, as BMI increases the effect of this term in leptin production also increases. The positive correlation of insulin in leptin concentration is modelled following Michaelis–Menten (MM) kinetics (represented as *T*_5_). The term *T*_6_ models leptin degradation. The newly introduced model parameters and some of the existing parameters are estimated using available data. This model (Eqns. 3-7) is considered as *leptin* model in this paper.

## 4 Parameter Estimation

The parameters of the augmented model are estimated using the dataset described in section 4.1. Seven new parameters - *g*_*l*_, *k*_*l*_, *k*_*b*_, *b*_*m*_, *k*_5_, *k*_6_ and *k*_*i*_ are introduced in the augmented model. Model fitting is done to minimise the error between the observed data and simulated behavior of the model.

### 4.1 Dataset used for parameter estimation

Two sets of data are used in the study. Dataset-I consists of 40 rows of data related to subjects with T2DM, collected from a community-based primary health clinic located in the district of North 24 Parganas of West Bengal, India, who underwent OGTT. After pre-processing (removal of rows with missing values) the dataset consisted of data from 38 subjects. The subjects were asked to ingest 75g of glucose dissolved in 100mL of water after fasting for 8-12 hours. The sample rows from dataset-I are shown in Table 1a. The dataset consists of the following columns: BMI, measurements of insulin, glucose and leptin at various time points. FGLU indicates fasting glucose level. GLU⟨*t*⟩, INS⟨*t*⟩ and LEP⟨*t*⟩ indicate glucose, insulin and leptin levels after time ⟨*t*⟩ of orally ingesting glucose beyond fasting glucose measurement (at *t* = 0), respectively. For example, GLU30 indicates glucose level after 30 min and LEP45 indicates leptin level after 45 min. The measurements are taken at times: 0, 15, 30, 45, 60, 75, 90, and 120 minutes. All participants gave informed consent and the study was approved by the Institutional Human Ethics Committee of CSIR-IICB.

**Table 1:**
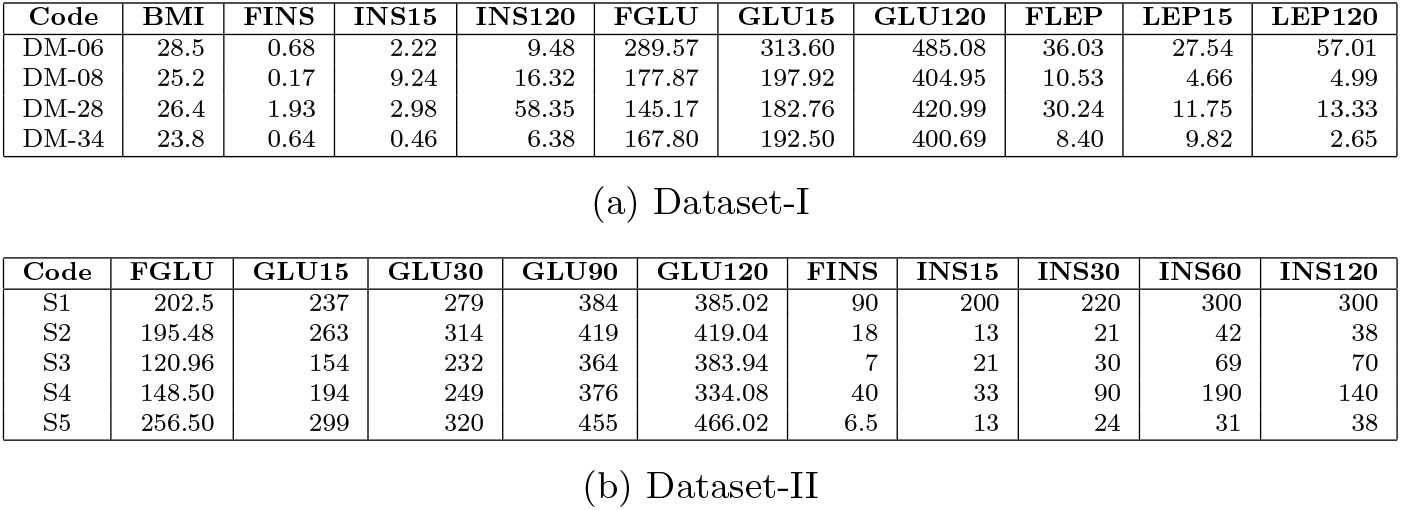
Sample rows from dataset-I and dataset-II. Glucose units are in *mg/dL* and insulin units are in *µU/mL* and.

Dataset-II consists of OGTT data of 129 individuals diagnosed with T2DM, collected from local pathology labs by a collaborator from a research institute. The subjects were asked to ingest 25g of glucose dissolved in 100mL of water after fasting for 8-12 hours. Sample data from dataset-I are given in Table 1b. The column names are similar to dataset-I with only changes in time points. The measurements in dataset-II are taken at times: 0, 15, 30, 60, 90, and 120 minutes. These data were obtained from local pathology laboratories in anonymized form, ensuring the protection of patient confidentiality. The dataset-I is used for estimation of parameters in the leptin model and dataset-II is used in validation of the model.

The condition which is characterized by abnormally high levels of insulin in the bloodstream (hyperinsulinemia) strongly influences leptin levels [6]. This is not accommodated in the present model as the available data used in the study only included subjects who were not affected by hyperinsulinemia.

### 4.2 Estimation Techniques

Parameter estimation is performed by solving an optimisation problem to minimise the squared error between outputs obtained from solving ODE and the observed data. The methods are implemented and coded in Python framework using the SciPy packages odeint() [36] and lmfit() [37]. Parameter estimation runs were performed for each individual subject, and average parameter values were calculated from the estimated results. Different estimation techniques were tried out, and Nelder-Mead optimisation [26] gave a better fit of observed data. The error function used in estimation is defined as follows:

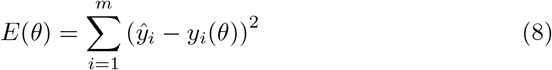

Where 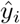 are observed data values and *y*_*i*_ are simulated values for a given parameter *θ* at *m* time points.

When performing parameter estimation or fitting for the system of ODEs, it is common to impose bounds on the parameters to ensure that the estimated values are within a reasonable and physiologically meaningful range. Setting parameter bounds helps to constrain the optimization process and prevents unrealistic or unbounded parameter estimates. This model considers parameter ranges reported in the relevant literature and physiological constraints while setting parameter bounds.

### 4.3 Parameter Sensitivity Analysis

Parameter sensitivity analysis is used to examine how sensitive a mathematical model responds to variations in its input variables. When every parameter in an ODE model is varied to understand the model response, global sensitivity analysis is used. The general formulation of a non-linear dynamic system is shown as:

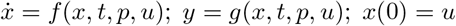

Where 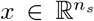 is the vector of *n*_*s*_ number of state variables, 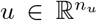 is the vector of *n*_*u*_ number of initial values of state variables, 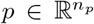 is the parameter vector of length *n*_*p*_ and 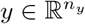 is the output vector of length *n*_*y*_. The first-order sensitivity function is a sampling time dependent matrix *S*_*it*_(*t*_*k*_) such that the sensitivity value of *i*^*th*^ output variable concerning *j*^*th*^ parameter at time *t*_*k*_ is defined as:

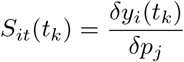

Parameter ranges for new parameters in the model are determined through sensitivity analysis. This is done by systematically varying the values of parameters within certain ranges and observing the impact on model behaviour. In this way, sensitivity analysis helps identify the ranges of parameter values that produce reasonable model responses constrained within physiologically plausible ranges.

## 5 Revision of indices derived from OGTT

Introducing a revised version of HOMA-B, HOMA2, and HOMA-IR, which incorporates BMI and fasting leptin, is a valuable approach to account for the influence of obesity on beta-cell function and insulin resistance. The resulting indices can provide a more comprehensive assessment of insulin secretion and sensitivity. HOMA-B focuses only on fasting glucose and insulin values, while DI_o_ also considers post-prandial values. The revised indices for HOMA-B, HOMA2, and HOMA-IR are represented as functions of BMI and leptin, as shown in the equations:

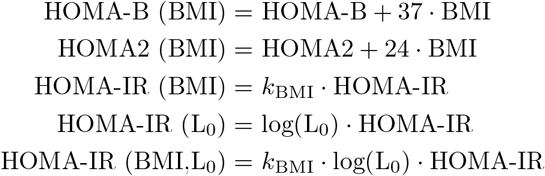

where L_0_ is fasting leptin in *ng/mL*. In order to establish a closer association with ISI, BMI is incorporated into HOMA-B and HOMA2 calculations by applying a scaling factor, and the resulting values are analyzed to determine their correlation with ISI. Through a systematic evaluation of scaling factor values, the optimal scaling factor, which exhibits the highest correlation with the ISI, is identified (as shown in Fig. 3). This iterative approach ensures that the scaling factor chosen is the minimum value that maximizes the required correlation. The HOMA-IR index is enhanced by incorporating *k*_BMI_ and L_0_ as separate variables, as well as by considering their combined effect. The correlation between the augmented HOMA-IR index and DI_o_ is also measured in each case to assess their relationship. By incorporating BMI and fasting leptin values into these indices, clinicians and researchers can gain insights into the combined effects of insulin secretion, insulin resistance, and obesity on T2DM.

**Fig. 3:**
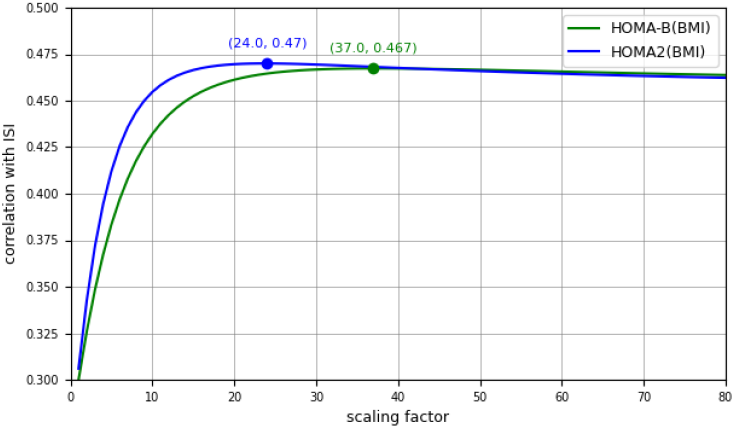
Variation of scaling factor used in revised HOMA-B and HOMA2 wrt correlation with ISI

## 6 Results and validation

This section presents the parameter estimation results of the leptin model, along with its analysis and validation. The estimated parameters provide valuable insights into the dynamics of glucose, insulin, and leptin concentrations in IG regulatory system, and validation with data ensures the reliability of *leptin* model.

### Results of Parameter Estimation

The model is estimated for parameter values and the resulting simulation closely matches with the observed data. A total of nine parameters are estimated out of the 24-time point data for each subject. Five new parameters - *g*_*l*_, *k*_*l*_, *k*_*b*_, *k*_6_ and *k*_*i*_ of *leptin* model and four parameters - *k*_1_, *k*_2_, *k*_3_ and *k*_4_ of the *base* model are estimated using the Nelder-Mead optimisation method. The parameter ranges are determined based on sensitivity analysis described in section 4. The parameter description, values, coefficient of variation (CV), and units are described in Table 2. The parameters of the *R*_*a*_ model for glucose absorption, *k*_*max*_, *k*_*min*_, *k*_*abs*_ and *f* are taken from literature[13]. Few parameters of the *leptin* model are calculated based on known relationships or processes. As BMI is usually in the range 18-32, the parameter *b*_*m*_ in *k*_BMI_ (Eqn 6) is fixed at 21 as the short range of the logistic function gives the effect of BMI adequately. This is the calculated average value of estimated *b*_*m*_ across all the subjects. The parameter *k*_5_, which is the rate constant for leptin degradation, is found to have a half-life of 26 min [35], and the value is fixed at 0.03. The simulation results of two sample subjects are shown in Fig. 4. The *leptin* model captures the observed time point measurements of leptin along with insulin and glucose. The model effectively captures the relationship between insulin and leptin, wherein an increase in insulin level leads to a decrease in leptin level. This agreement between the model predictions and observed behaviour highlights the ability of the model to represent the regulation between insulin and leptin accurately. CV is calculated to express the relative variability in the parameters. For instance, a CV value of *k*_*b*_ exhibits less variability and more precision, whereas *k*_*l*_ indicates a higher CV which attributes to the range of BMI in subjects (Table 2).

**Table 2:** Estimated parameters in *leptin* model.

| Symbol | Description | Value | CV | Source | Unit |
| --- | --- | --- | --- | --- | --- |
| $g_l$ | the ability of leptin to lower blood glucose levels | 3.66 | 2.85 | Estimated | $\frac{Lmin^{-1}}{\mu mol}$ |
| $k_l$ | ability of insulin to stimulate leptin production | 4.99e-05 | 1.92 | Estimated | $min^{-1}$ |
| $k_5$ | leptin degradation rate | 0.03 | | Calculated from [35] | $min^{-1}$ |
| $k_6$ | the inhibitory action of leptin on insulin | 749.74 | 2.80 | Estimated | $L/\mu mol$ |
| $k_b$ | BMI contribution factor to leptin production | 9.41e-05 | 0.89 | Estimated | $min^{-1}$ |
| $k_i$ | leptin production based on insulin | 5.15e-05 | 4.15 | Estimated | $min^{-1}$ |
| $b_m$ | scaling factor that regulate effect of BMI | 21 | | Calculated from Eqn. 6 | constant |
| $k_1$ | glucose effectiveness; the ability to promote glucose disposal and inhibit glucose production | 0.027 | | Estimated | $min^{-1}$ |
| $k_2$ | the insulin-dependent increase in glucose uptake ability in tissue per unit of insulin concentration above $I_b$ | 0.110 | | Estimated | $min^{-1}$ |
| $k_3$ | scale factor for the amplitude of insulin action | 7.634 | | Estimated | $\frac{min^{-2}}{(\mu mol/L)^{-1}}$ |
| $k_4$ | decay rate for insulin in plasma | 3.349 | | Estimated | $min^{-1}$ |
| $\gamma$ | the rate of the pancreatic β-cells' release of insulin after the glucose injection and with glucose concentration above $D$ | 0.017e-6 | | Estimated from base model | $\mu mol/L$ |

**Fig. 4:**
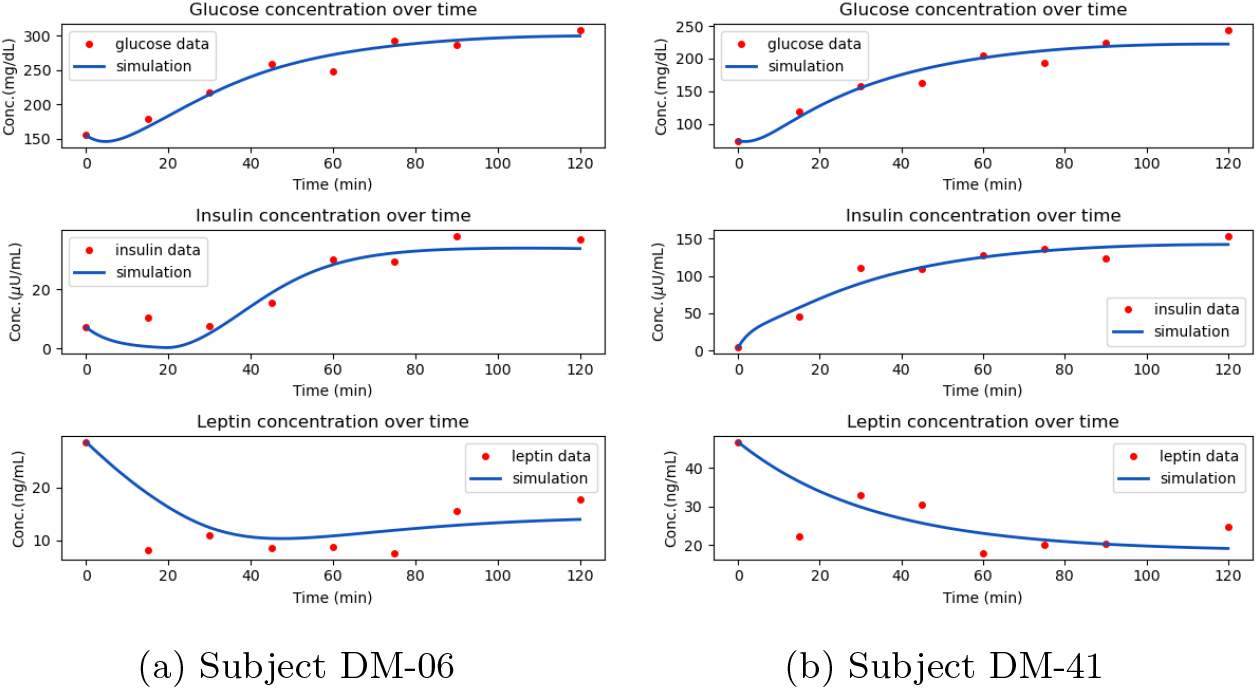
Simulation results sample showing glucose, insulin and leptin concentrations for subjects

**Fig. 5:**
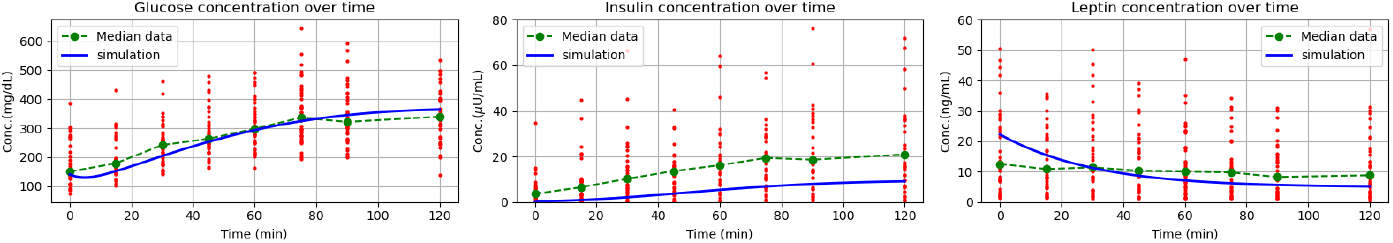
Glucose, insulin, and leptin concentration range for all study participants (n=38) for 2-h OGTT. Median glucose, insulin, and leptin concentrations from data (dashed green line) and simulation of *leptin* model (blue line) for a BMI = 25.

The timepoint measures of glucose, insulin and leptin from dataset-II, along with their median values, are shown in Fig. 5. The simulation result of *leptin* model is also shown to compare it with the median data. In the insulin graph, the simulation shift observed in the farther time points are within the variation of insulin, and for glucose and leptin, the variation is not profound

### Results of Parameter Sensitivity Analysis

The newly introduced parameters are sensitive to the observables highlighting the positive aspects of the current model. Sensitivity analysis is done on five parameters *k*_*l*_, *k*_*b*_, *g*_*l*_, *k*_6_ and *k*_*i*_ in *leptin* model, considering a ± 30% variation with respect to glucose, insulin, and leptin, and the results are shown in Fig. 2. The parameter *g*_*l*_ is highly sensitive to glucose and insulin whereas less sensitive to leptin which implies the ability of leptin to lower blood glucose levels impacts model behavior significantly. This variation in *g*_*l*_ analysis helps in incorporating adequate ranges for *g*_*l*_. Insulin concentration is affected by *k*_6_, and is moderately sensitive to the inhibitory action of leptin on insulin. *k*_*b*_ is highly sensitive to insulin, glucose, and leptin, emphasizing the role of BMI in the *leptin* model. *k*_*l*_ and *k*_*i*_ are related to the insulin action in the production of leptin and are less sensitive to insulin giving only a small boost in insulin concentration.

### Validation of the augmented model and parameters

To evaluate whether the developed *leptin* model is capable of capturing the underlying dynamics of the insulin-glucose system, model validation is performed. The newly introduced parameter values are taken from the augmented model, and simulations are run for all the subjects. The BMI value is made a parameter for this simulation, and the estimated average is 24.6. Other parameter values are taken from literature as given in Table 2. The simulations performed with the *leptin* model demonstrated a better fit to the observed data points when compared to the *base* model. Sample simulation results of the *base* model and *leptin* model using dataset-II are shown in Fig 6. The simulated behavior of leptin is obtained, revealing a pattern that closely resembles the results previously observed for dataset-I. To assess the statistical significance of the differences, a Chi-square goodness of fit test (*α* = 0.05) was performed on the simulated and observed data. The analysis revealed a statistically significant result of p-values with 74% good fits using the *leptin* model compared to 62% good fits using the *base* model.

**Fig. 6:**
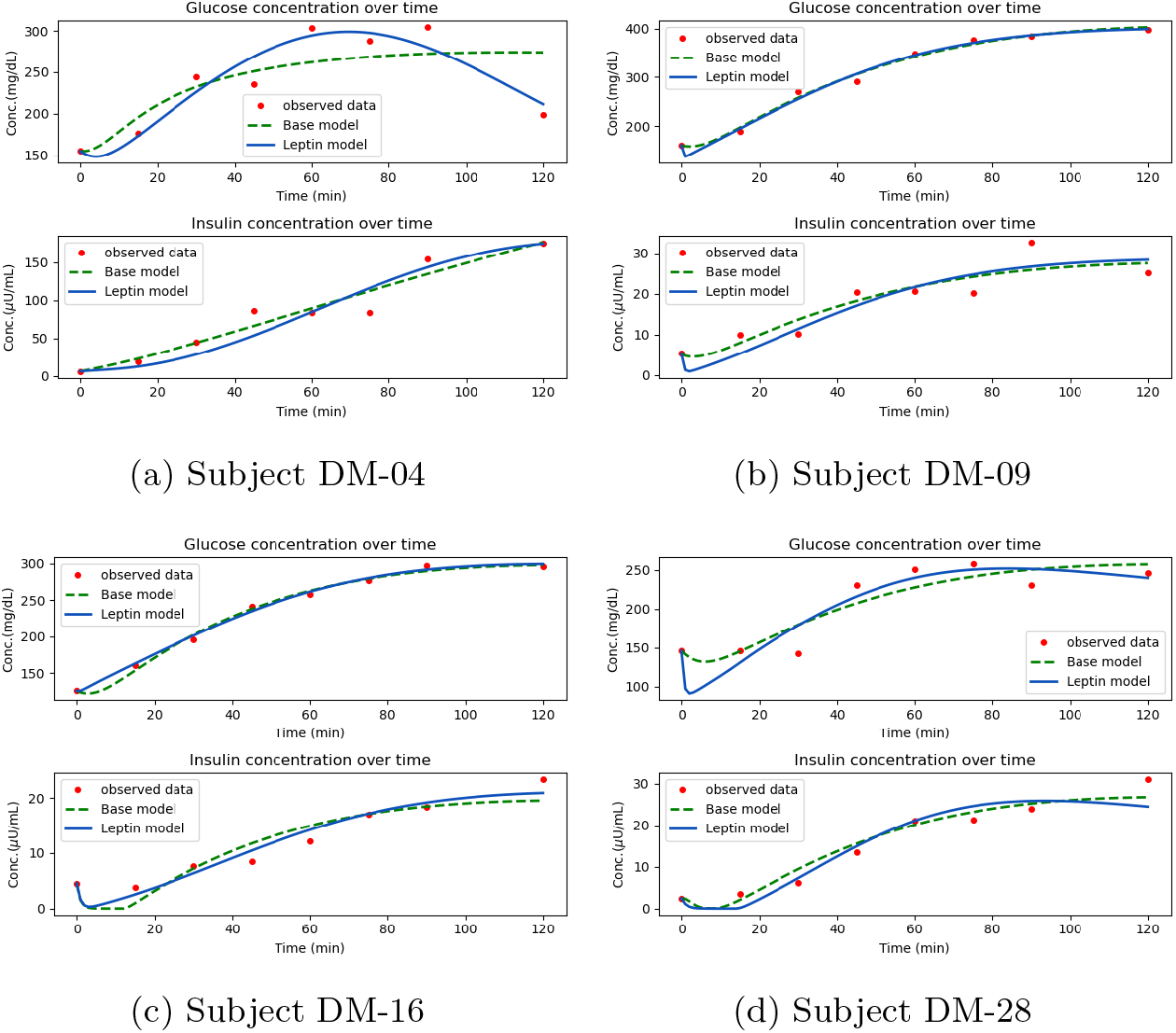
Plots of sample subjects showing glucose and insulin (for 75g glucose) for *base* model and *leptin* model using dataset-I

The *leptin* model is also validated using dataset-I without considering leptin values and the fittings were found to be better. A sample simulation results of the *base* model and *leptin* model using dataset-I are shown in Fig 7.

**Fig. 7:**
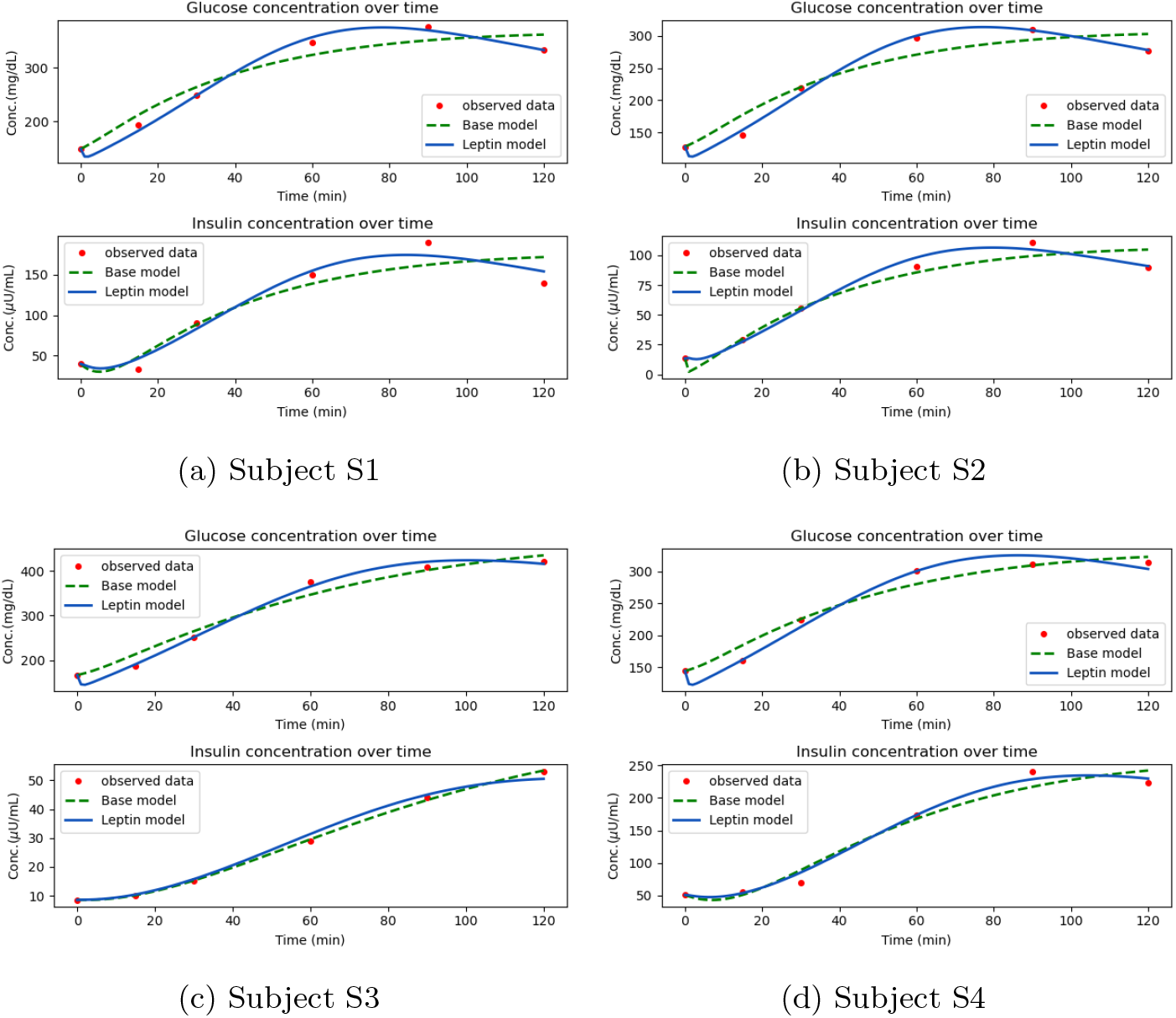
Plots of sample subjects showing glucose and insulin (for 25g glucose) for *base* model and *leptin* model using dataset-II

### Results of revised OGTT indices

The correlation values between indices are shown in Table. 3. The inclusion of BMI in HOMA-B and HOMA2 exhibits a better correlation with ISI with coefficients of 0.467 and 0.469, respectively. This observation suggests that the inclusion of BMI enhances the indices by establishing a closer association with ISI.

**Table 3:**
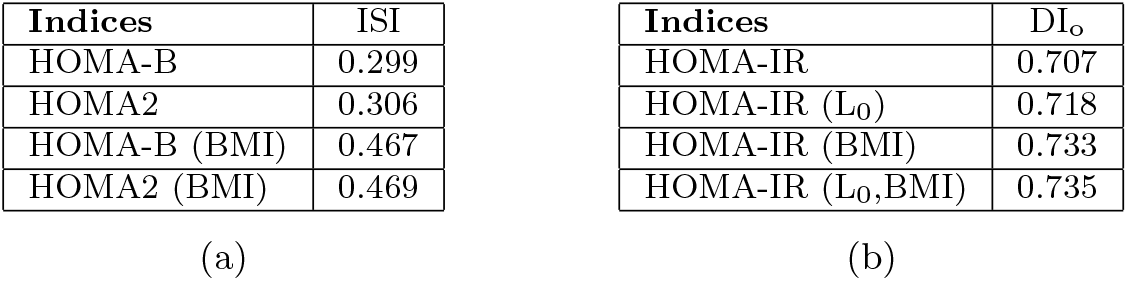
(a) Correlation of ISI to HOMA-B and HOMA2 and their revisions (b) Correlation of DI_o_ to HOMA-IR and revised HOMA-IR.

When examining the correlation between HOMA-IR and DI, the inclusion of BMI and leptin as separate variables, as well as their combined effect, resulted in slightly higher correlation values compared to the existing correlation between these factors. This suggests that BMI and leptin contribute to the relationship between HOMA-IR and DI, and their combined impact further strengthens their association. These revised indices may be used clinically for better assessment of *β*-cell function and insulin resistance.

## 7 Discussion

The objective of this work is to augment the existing mathematical model of IG regulatory system to accommodate the role of leptin and BMI. Parameters for the *leptin* model are estimated using the observed data of glucose, insulin and leptin for individuals with T2DM. The simulating results and the observed data have a statistically significant Chi-square fit of 74%. As leptin is involved in glucoregulatory actions, predicting its behaviour through the *leptin* model can help in developing targeted interventions and therapies for managing and treating T2DM taking into account obesity-related issues. By influencing insulin secretion and sensitivity, leptin affects glucose metabolism and overall glycemic control in individuals with T2DM. Leptin is reported to have potent anti-diabetic actions independently of its effects on body weight and food intake [38]. These anti-diabetic effects of leptin can be explored and used in leptin therapy for the treatment of T2DM [38]. Further, longterm leptin-replacement therapy can significantly improve glycemic control and insulin sensitivity in patients with severe IR [39]. The new model can predict leptin behaviour in association with insulin and glucose concentrations using clinically significant parameters. A personalised treatment approach by integrating leptin behaviour may lead to more effective and targeted treatments for metabolic conditions.

Revised OGTT indices (HOMA-B (BMI), HOMA2 (BMI), HOMA-IR (BMI), HOMA-IR (L_0_), HOMA-IR (BMI, L_0_)) have been proposed. These can aid in better insulin dosage estimation. The effect of the estimated dosage may be better predicted using the *leptin* model. T2DM therapy largely depends on the degree of IR and insulin secretory dysfunction, the contribution of which is associated with obesity. Hence, the inclusion of obesity parameters in assessing IR and *β*-cell dysfunction is vital in the decision-making process of anti-diabetic therapy. The simple and usable indices proposed in this work will help decide a precision anti-diabetic therapy for non-obese T2DM patients and may open the door to other possibilities to build more accurate obesity-associated indices for personalised medicine. The claims in the work are based on the available data of 38 subjects. It would be desirable to perform the study on larger data to increase the confidence of the results. A potential future work may be to enhance the presented *leptin* model by incorporating other adipokines (such as adipsin, adiponectin) that also play a role in the insulin-glucose regulatory system.

## 8 Conclusion

In this study, the OMM is augmented by incorporating the influence of BMI and leptin levels in the insulin-glucose regulatory system. The model is developed based on several theoretical studies on the association of leptin with insulin and glucose concentrations. The model parameters are estimated and analyzed using the OGTT data collected in a clinical setting. The new model offers valuable insights into the working of the insulin-glucose regulatory system taking into account the obesity of the subject and uncovering related risk factors involved in T2DM. This seems to be the first attempt to model the role of obesity indicators leptin and BMI in glucose regulation using ordinary differential equations (ODE). The revised OGTT indices, with the inclusion of obesity parameters, hold the potential to aid in determining precise antidiabetic therapies for T2DM. Simulation results from the augmented model with the identified parameters have a good match with the observed data of insulin, glucose, and leptin. The results presented in this work are based on limited available data. It would be desirable to reinforce those by extending the study to larger data.

## Data Availability

All data produced in the present study are available upon reasonable request to the authors

## Acknowledgement

The authors gratefully thank the associates of K V Venkatesh for collecting the insulin-glucose time series data. This work was partly funded by MoE, Government of India.

